# Data-driven estimation of the instantaneous reproduction number and growth rates for the 2022 monkeypox outbreak in Europe

**DOI:** 10.1101/2022.12.20.22283714

**Authors:** Fernando Saldaña, Maria L. Daza-Torres, Maíra Aguiar

## Abstract

**Objective:** To estimate the instantaneous reproduction number *R*_*t*_ and the epidemic growth rates for the 2022 monkeypox outbreaks in the European region.

**Methods:** We gathered daily laboratory-confirmed monkeypox cases in the most affected European countries from the beginning of the outbreak to September 23, 2022. A data-driven estimation of the instantaneous reproduction number is obtained using a novel filtering type Bayesian inference. A phenomenological growth model coupled with a Bayesian sequential approach to update forecasts over time is used to obtain time-dependent growth rates in several countries.

**Results:** The instantaneous reproduction number *R*_*t*_ for the laboratory-confirmed MPX cases in Spain, France, Germany, the UK, the Netherlands, Portugal, and Italy. At the early phase of the outbreak, our estimation for *R*_*t*_, which can be used as a proxy for the basic reproduction number *R*_0_, was 2.32 (95% CI 1.81 *−* 3.05) for Spain, 2.91 (95% CI 2.33 *−* 3.52) for France, 1.84 (95% CI 1.55 *−* 2.31) for UK, 3.16 (95% CI 2.55 *−* 3.64) for Germany, 2.97 (95% CI 2.01 *−* 4.32) for the Netherlands, 1.18 (95% CI 0.98*−*1.33) for Portugal, 3.74 (95% CI 2.91*−*4.49) for Italy. Cumulative cases for these countries present subexponential rather than exponential growth dynamics.

**Conclusions:** Our findings suggest that the current monkeypox outbreaks present limited transmission chains of human-to-human secondary infection so the possibility of a huge pandemic is very low. Confirmed monkeypox cases are decreasing significantly in the European region, the decline might be attributed to public health interventions and behavioral changes in the population due to increased risk perception. Nevertheless, further strategies toward elimination are essential to avoid the subsequent evolution of the monkeypox virus that can result in new outbreaks.

## 1 Introduction

The monkeypox virus (MPXV) is an enveloped double-stranded DNA virus discovered in a Danish laboratory in 1958 and is known for being the etiological agent of a zoonotic disease known as monkeypox (MPX) whose first human cases were identified in the Democratic Republic of Congo in 1970. Considering genetic and geographic variation, there are two distinct clades of the virus: the central African (also known as Congo Basin) and the west African clade, the former being the more virulent based on a higher case fatality ratio Poland et al. (2022). Since 1970, MPX has remained endemic in central and west Africa, with only a few cases outside of this region. Yet, in 2003 an outbreak was documented in the United States of America after the importation of infected animals. Until May 2022, all laboratory-confirmed MPX cases identified outside of West and Central Africa were either imported or linked to an imported case or animals imported from endemic areas Poland et al. (2022). On 16 May 2022, the United Kingdom Health Security Agency reported the identification of four cases of MPX with no history of recent travel to endemic areas or contact with previously reported cases. Afterward, multiple MPX outbreaks were reported in several countries. Hence, on 23 July, the WHO declared the outbreak a public health emergency of international concern World Health Organization (WHO) (2022). The outbreak has now affected more than 100 countries and more than 80, 000 cases have been confirmed as of 14 November 2022. Europe was the epicenter of this large and geographically widespread outbreak approximately until the end of July 2022. Later, the region of the Americas became the area with the highest number of confirmed cases World Health Organization (WHO) (2022).

People can not only acquire the MPXV after being in contact with an infected human but also via a spillover event after direct contact with body fluids, skin, or mucosal lesions of infected wildlife. Rodents, squirrels, non-human primates, and other species can be infected by MPXV, but the natural reservoir has not yet been confirmed. Human-to-human transmission occurs mainly through close contact with infectious material from skin lesions of an infected person and respiratory droplets after prolonged close contact Poland et al. (2022). The present outbreak has affected predominantly the gay and bisexual men who have sex with men (GBMSM) community, with the nature of the presenting lesions suggesting that transmission occurred during sexual intercourse, although some cases have also been detected in women and children World Health Organization (WHO) (2022). Mass gatherings events such as summer music festivals and specific sexual practices have facilitated the transmission of MPX among the GBMSM group European Centre for Disease Prevention and Control (2022). However, at present, there is not enough evidence to confirm that MPX transmission occurs mainly via sexual routes Zumla et al. (2022). Furthermore, while detecting and preventing human-to-human secondary infections are critical, transmission from animals to humans is also a major concern. If the virus becomes endemic in an animal reservoir outside of endemic areas, it would mean a continuous risk of repeated human outbreaks, and disease elimination will not be reachable Poland et al. (2022).

As the monkeypox outbreak unfolds, mathematical modeling can be used as a public health guidance tool to evaluate the impact of control interventions and better understand the epidemiology of the outbreak Yuan et al. (2022). Real-time monitoring of changes in transmissibility can be done by virtue of the instantaneous reproduction number *R*_*t*_ (also called effective reproduction number), defined as the mean number of infections produced by a typical infectious case at calendar time *t* Saldaña and Velasco-Hernández (2022). A reliable estimation of R_*t*_ is useful to quantify if the outbreak is declining (*R*_*t*_ *<* 1), growing (*R*_*t*_ *>* 1), or plateauing (*R*_*t*_ *<* 1), and can be used to evaluate and adjust health interventions in real time Parag et al. (2022). A classical assumption in epidemic modeling is that in the early phase when susceptible depletion is negligible or control measures are absent, outbreaks show exponential growth dynamics. Nevertheless, early subexponential growth dynamics have already been documented in empirical data for multiple disease outbreaks including HIV and Ebola virus Chowell et al. (2016); Chowell (2017); Viboud et al. (2016). The mechanisms that lead to subexponential growth profiles are still debated but include heterogeneity in contact structures, clustering of contacts, and individual behavioral changes due to an increased risk perception Chowell et al. (2016). These mechanisms have been present in the current outbreaks Poland et al. (2022); Zumla et al. (2022). Hence, we consider the possibility of early subexponential growth dynamics on the MPX outbreaks using a flexible phenomenological model that is able to reproduce several growth profiles Chowell et al. (2016).

The main goal of this work is to obtain a data-driven estimation of *R*_*t*_ and growth rates for the MPX outbreak in the most affected European countries using incidence time series based on confirmed cases. To obtain a reliable estimation of the instantaneous reproduction number we use a recently proposed filtering type Bayesian inference Capistrán et al. (2022). This estimation considers the mean generation time for the current MPX outbreak. Furthermore, we pose a novel adaptive scheme to consider non-autonomous epidemiological dynamics, in particular, time-dependent dynamics of the intrinsic epidemic growth rate in a generalized growth model. To this end, the number of MPX cases is modeled through a sequential Bayesian approach Daza-Torres et al. (2022) with a Negative Binomial (NB) model.

## 2 Methods and Results

### 2.1 The data

According to the European Center for Disease Control (ECDC), as of November 22, 2022, a total of 17,395 MPX cases have been confirmed in Europe. Spain (7,405), France (4,104), United Kingdom (3,720), Germany (3,672), the Netherlands (1248), Portugal (948), and Italy (917) are the most affected countries in terms of confirmed cases within the European region European Centre for Disease Prevention and Control (2022). We present daily numbers of laboratory-confirmed MPX cases from the start of the outbreak in each country, respectively, until 23 September 2022 (see Figure 1). Following this period most of these countries report very few cases for several weeks. To account for reporting delays and other incidence data anomalies, a 10-day moving average of cases is also presented (see Figure 1). The incidence time series and confirmed cases for Europe and several other countries can be obtained from an open-access database presented in Kraemer et al. (2022). In the case of the European Region, data is also available via the European Center for Disease Control (ECDC) and the WHO Regional Office for Europe through The European Surveillance System (TESSy) European Centre for Disease Prevention and Control (2022). Using the confirmed daily cases we obtained the cumulative cases in the European countries with the highest disease burden (see Figure 2).

**Figure 1:**
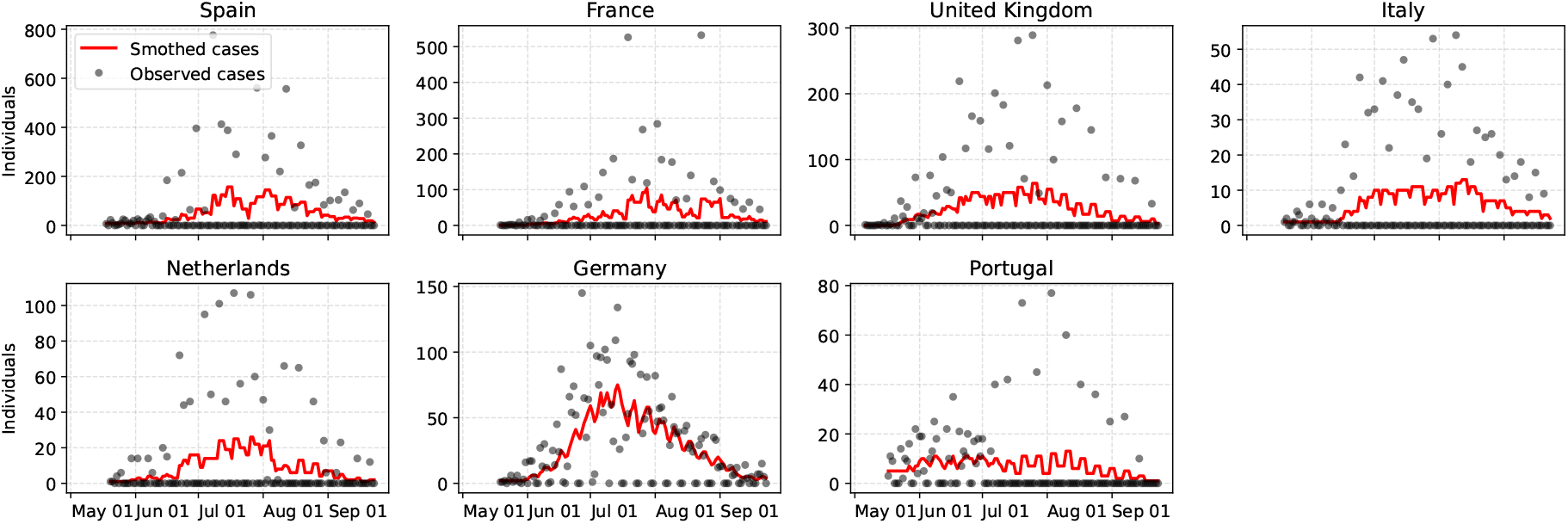
Daily laboratory-confirmed monkeypox cases in Spain, France, the UK, Italy, the Netherlands, Germany, and Portugal since the beginning of the outbreak to September 23, 2022. Raw incidence counts are presented as points and a 10-day moving average of (smoothed) cases is shown via a solid red line.

**Figure 2:**
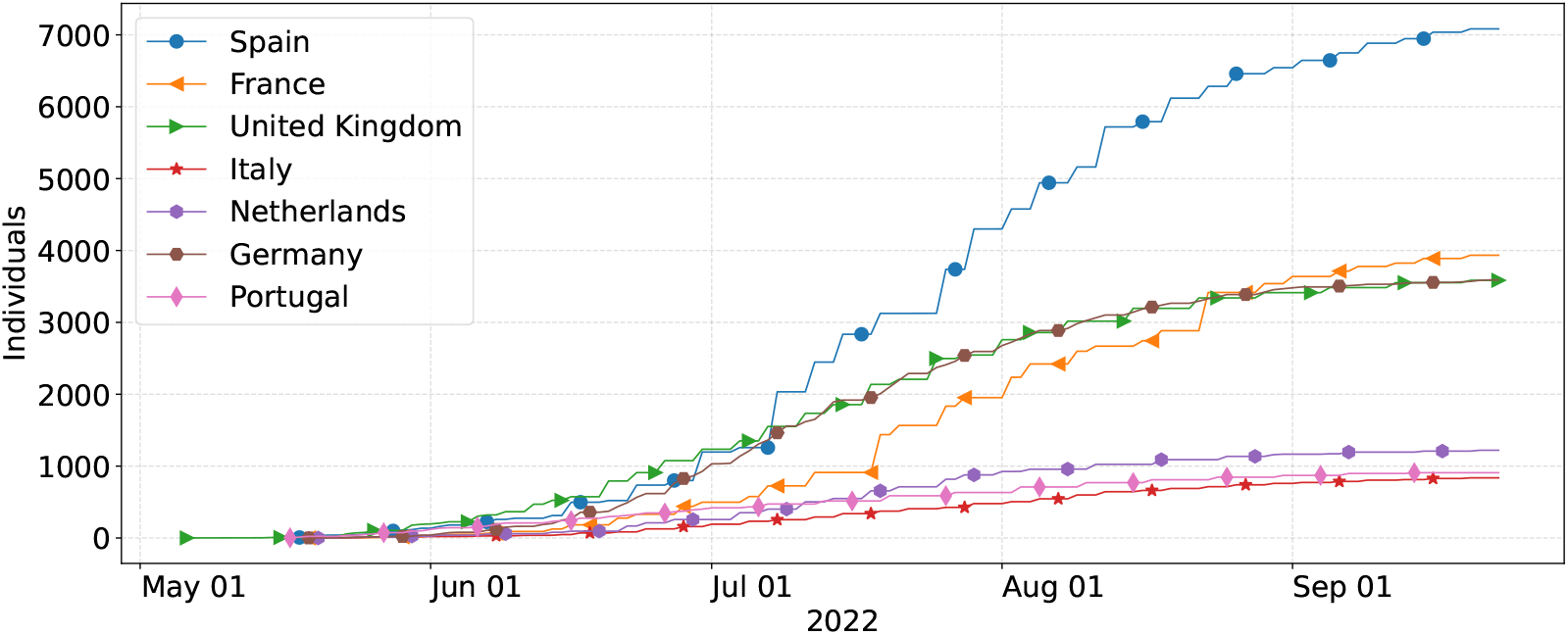
The cumulative number of confirmed monkeypox cases in Spain, France, Germany, the UK, the Netherlands, Portugal, and Italy from the beginning of the outbreak to September 23, 2022.

### 2.2 Basic and instantaneous reproduction numbers

The basic reproduction number, *R*_0_, is a central concept in the modeling of infectious diseases. *R*_0_ is defined as the expected number of secondary infections produced by a typical infected individual in a population where everyone is susceptible. Observe that *R*_0_ considers a fully susceptible population and, hence, control measures or behavioral changes technically do not reduce its value. In the presence of time-dependent changes in the population, the instantaneous reproduction number, *R*_*t*_, which is defined as the average number of secondary cases produced by one infected individual at time *t* is a key metric for evaluating the transmissibility of infectious diseases. Both reproduction numbers have been used successfully to monitor contagion patterns in past epidemics including the COVID-19 pandemic Aguiar et al. (2020); Saldaña and Velasco-Hernández (2021). Several methods have been developed to estimate both reproduction numbers. For example, the basic reproduction number can be expressed as Wallinga and Lipsitch (2007)

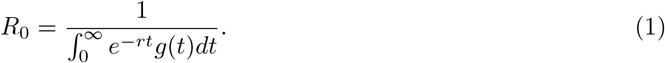

Here, the exponential growth rate, *r*, is defined as the per capita change in the number of new infections per unit of time. The function *g*(*t*) is the density of the generation interval *G* which is the time lag between infection in a primary case and a secondary case. The generation time distribution *g*(*t*) should be estimated empirically by considering the time lag between confirmed infector-infectee pairs. Nevertheless, in practice, it is difficult to obtain and is commonly substituted with the serial interval distribution that measures the time from illness onset in the primary case to illness onset in the secondary case Obadia et al. (2012). Several methods have been proposed to estimate *R*_*t*_ using only incidence data e.g. Capistrán et al. (2022); Cori et al. (2013); Fraser (2007); Obadia et al. (2012). Using the renewal equation Fraser (2007), the instantaneous reproduction number can be expressed in terms of the incidence *I*_*t*_ at time *t*, and the discretized probability distribution of the generation interval denoted *g*_*s*_ as follows

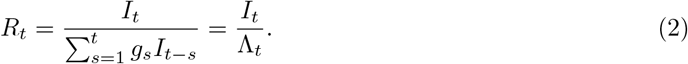

The denominator Λ_*t*_ is usually interpreted as the total infectiousness of infected individuals Cori et al. (2013). Furthermore, Λ_*t*_ can also represent an estimation of the current number of active cases, so *R*_*t*_ is the ratio of secondary cases produced by the actual total active cases Capistrán et al. (2022). Given the definition (2), Cori et al. (2013) assumed that

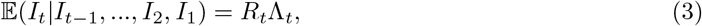

where E(*X*) denotes the expectation of a random variable *X*, and *I*_*t*_ conditional on previous incidences follows a Poisson model i.e. *I*_*t*_| *I*_*t−*1_, …, *I*_2_, *I*_1_ *∼Po*(*R*_*t*_Λ_*t*_). Nevertheless, overdispersion is expected for contagious events, so a natural improvement on the Poisson model would be to consider a more general count data model such as a Negative Binomial. Here, we use a recently proposed method Capistrán et al. (2022) to obtain a reliable estimation of *R*_*t*_ that improves the Poisson sampling model by adding an autoregressive prior for the log of observed *R*_*t*_’s. This results in a dynamic linear model which coupled with Bayesian updating forms a filtering type inference for the sequence of *log*(*R*_*t*_) (see the details in Capistrán et al. (2022)).

One key input to calculate *R*_*t*_ is the generation time distribution. The mean generation time for the current MPX outbreak in Italy Guzzetta et al. (2022) was estimated to be 12.5 days (95% CI of the mean: 7.5*−*17.3 and 95^*th*^ percentiles of the distribution: 4*−* 26). We postulate a Gamma distribution with scale 0.5 and shape parameter 25, *Gamma*(25, 0.5), depicted in Figure 3 to approximate the generation time distribution and assume that this estimate for Italy is valid for the European region. Considering the possible presence of noise due to delays in reporting and other incidence data anomalies in epidemiological data, a common practice is to consider a smoothed version, S[*I*_*t*_], of the incidence time series. Several methods are available to obtain S[*I*_*t*_] curves e.g. splines of moving average filters (see Parag et al. (2022) and the references therein). Since *R*_*t*_ estimations based on very low incidence numbers are unreliable Cori et al. (2013) and the MPX incidence time series present several days with zero cases, we consider the 10-day moving average for the daily cases presented in Figure 1.

**Figure 3:**
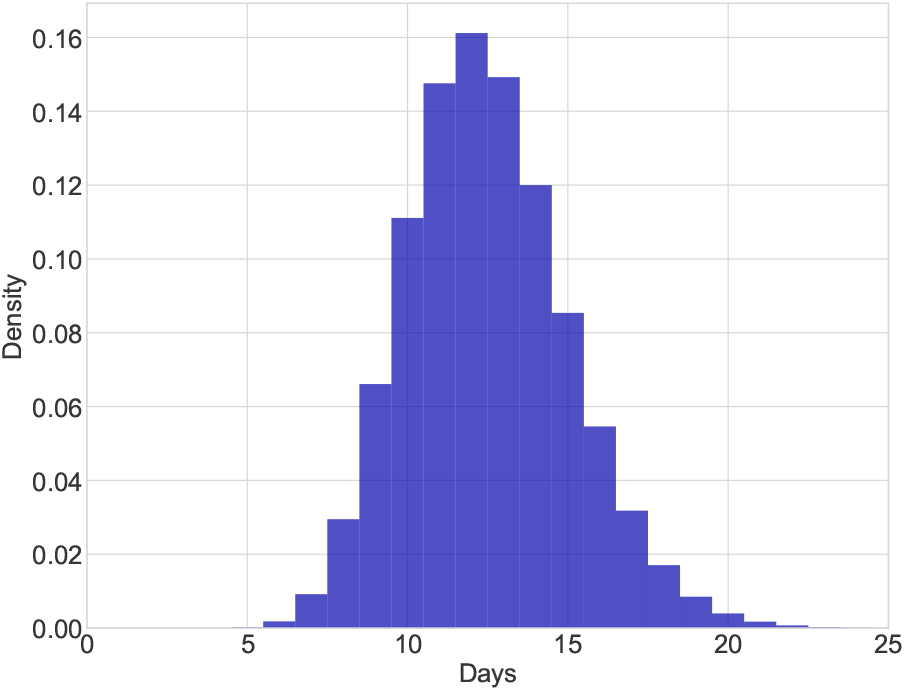
Histogram for the postulated *Gamma*(25, 0.5) that mimics current estimations for the generation time distribution of the 2022 MPX outbreaks.

### 2.3 Phenomenological models

Phenomenological models have been proven to be useful tools to characterize and generate shortterm forecasts of the evolution of epidemic outbreaks Chowell (2017). Phenomenological approaches prioritize the reproducibility of empirical observations. Hence, these models are particularly helpful in the presence of uncertainty of major epidemiological aspects including the potential contribution of multiple transmission pathways Chowell et al. (2016). This is the case for MPX and other zoonotic diseases for which not only human-to-human infection is possible but also cross-species transmission via spillover events. Key quantities such as the growth rate, peak date, the epidemic’s final size, and the epidemic wave’s length can be estimated via phenomenological models. The early dynamics of epidemic outbreaks are usually characterized by exponential or sub-exponential growth, followed by a deceleration of growth due to control measures or behavioral changes in the population. Several phenomenological models such as the generalized growth model (GGM), the Richards model (RM), and the Blumberg hyperlogistic function have been used successfully to generate a variety of epidemic growth profiles observed in real epidemic outbreaks Chowell (2017); Chowell et al. (2016); Viboud et al. (2016). Most of these models can be obtained from the generalized logistic growth model (GLGM) Tsoularis and Wallace (2002) defined as

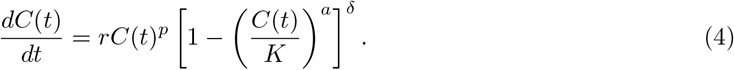

where *p, a*, and *δ* are non-negative real numbers.

Here, we consider the GGM model as a tool to characterize the epidemiological dynamics of the 2022 outbreak in the European region. The GGM is a particular case of the GLGM with *δ* = 0, and is defined by the solution of the following differential equation:

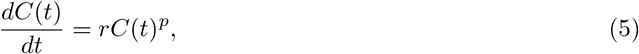

where *C*(*t*) represents the confirmed cumulative cases at time *t, r* is the growth rate, and *p* reflects the “deceleration of growth”. The value of *p* lies in the interval [0, 1] and allows the epidemic curve to mimic exponential (*p* = 1), sub-exponential (0 *< p <* 1), and linear growth (*p* = 0) Chowell (2017). The solution of the GGM can be easily obtained as

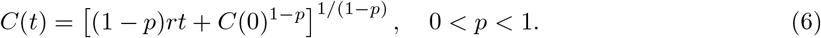

For *p* = 1, the solution is the classical exponential growth model *C*(*t*) = *C*(0)*e*^*rt*^. Whereas for *p* = 0, we have the linear growth solution *C*(*t*) = *rt* + *C*(0).

### 2.4 Bayesian parameter inference

We perform a simultaneous estimation of parameters *r* and *p* that focus on the epidemic growth phase of the MPX outbreaks. To this end, we adopt a Bayesian statistical approach, which is well suited to model multiple sources of uncertainty and allows the incorporation of background knowledge on the model’s parameters.

#### 2.4.1 Observational model and data

The observed data used to fit the model is based on daily laboratory-confirmed MPX cases (see Figure 1). We consider daily cases counts and their corresponding theoretical expectation *µ* at a discrete time *t*_*i*_, defined as

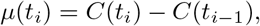

where *C*(*t*) is given by the subexponential solution (6) of the GGM model. We consider daily confirmed cases of patients with positive tests for the area being analyzed. To account for over-dispersed counts, we use a negative binomial (NB) distribution *NB*(*µ, ω, θ*) with mean *µ* and over-dispersion parameters *θ* and *ω*. For data *y*_*i*_, we let

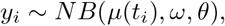

with fixed values for the over-dispersion parameters *ω, θ*. Conditional independence is assumed in the data and therefore the likelihood is obtained from the NB model.

The parameters to be inferred are the growth rate (*r*), the deceleration of growth (*p*), and we also infer the initial condition (*C*_0_). To sample from the posterior, we make use of Markov Chain Monte Carlo (MCMC) using the t-walk generic sampler Christen and Fox (2010). The MCMC runs semi-automatic, with consistent performances in most data sets.

#### 2.4.2 Bayesian sequential approach

Modeling studies often consider epidemiological dynamics as an autonomous dynamical system and neglect time-dependent changes in epidemiological parameters Hadeler (2011). Nevertheless, parameters usually evolve during the outbreak due to the impact of health interventions and changes in risk perception Camacho et al. (2019); Saldaña and Barradas (2019); Weitz et al. (2020). Here, we adapt the sequential data assimilation approach proposed in Daza-Torres et al. (2022) to obtain time-varying parameters. This approach assumes that suitable transmission, epidemic, and observation models are available and coded into a dynamical system. The main idea is to train the model using only a subset of the most recent data. The estimation is updated sequentially in a sliding window of data. We apply this method to understand the disease dynamic in the growth stage of the outbreak and track changes over the estimated parameters in the phenomenological model (6) using incidence time series for MPX in European countries.

Regarding the elicitation of the parameters’ prior distribution for the first forecast, we use a Gamma distribution for the initial condition, *C*_0_, with scale 1 and shape parameter 10. This assumption is appropriate to model low incidence counts, near to 10, on the initial number of infected individuals. The prior distributions for the remaining parameters are summarized in Table 1. We have to stress that prior distributions are only used at the first learning window. Then, the MCMC posterior sample from window *k* is used to create a prior for the next window *k* + 1 (see details in Daza-Torres et al. (2022)).

**Table 1:** Parameters and prior distributions for the first learning window used for Bayesian inference. The prior distributions presented here are only used at the start and are not used in the rest of the sequential inference, wherein each window, the prior is an over-dispersed version of the posterior in the previous window.

| Parameter | Prior distribution |
| --- | --- |
| Growth rate $r$ | $r \sim N(1, 1)$ |
| Deceleration of growth $p$ | $Beta(1 + 1/6, 1 + 1/3)$ |

## 3 Results

### 3.1 The instantaneous reproduction number

The instantaneous reproduction number *R*_*t*_ for the laboratory-confirmed MPX cases in Spain, France, Germany, the UK, the Netherlands, Portugal, and Italy from June 25, to September 23, 2022, is presented in Figure 4. We used a *Gamma*(25, 0.5) distribution (see Figure 3) as an approximation of the generation time distribution for the European region. In Figure 4, the red solid line represents the median estimate for *R*_*t*_, whereas the dark and light blue shaded areas represent 50% and 90% quantiles, respectively. The green solid line indicates the threshold value 1 on the reproduction number. At the beginning of the period, our estimation for *R*_*t*_, which can be used as a proxy for the basic reproduction number *R*_0_, was 2.32 (95% CI 1.81*−*3.05) for Spain, 2.91 (95% CI 2.33*−*3.52) for France, 3.16 (95% CI 2.55*−*3.64) for Germany, 1.84 (95% CI 1.55*−*2.31) for the UK, 2.97 (95% CI 2.01*−*4.32) for the Netherlands, 1.18 (95% CI 0.98*−*1.33) for Portugal, 3.74 (95% CI 2.91*−*4.49) for Italy.

**Figure 4:**
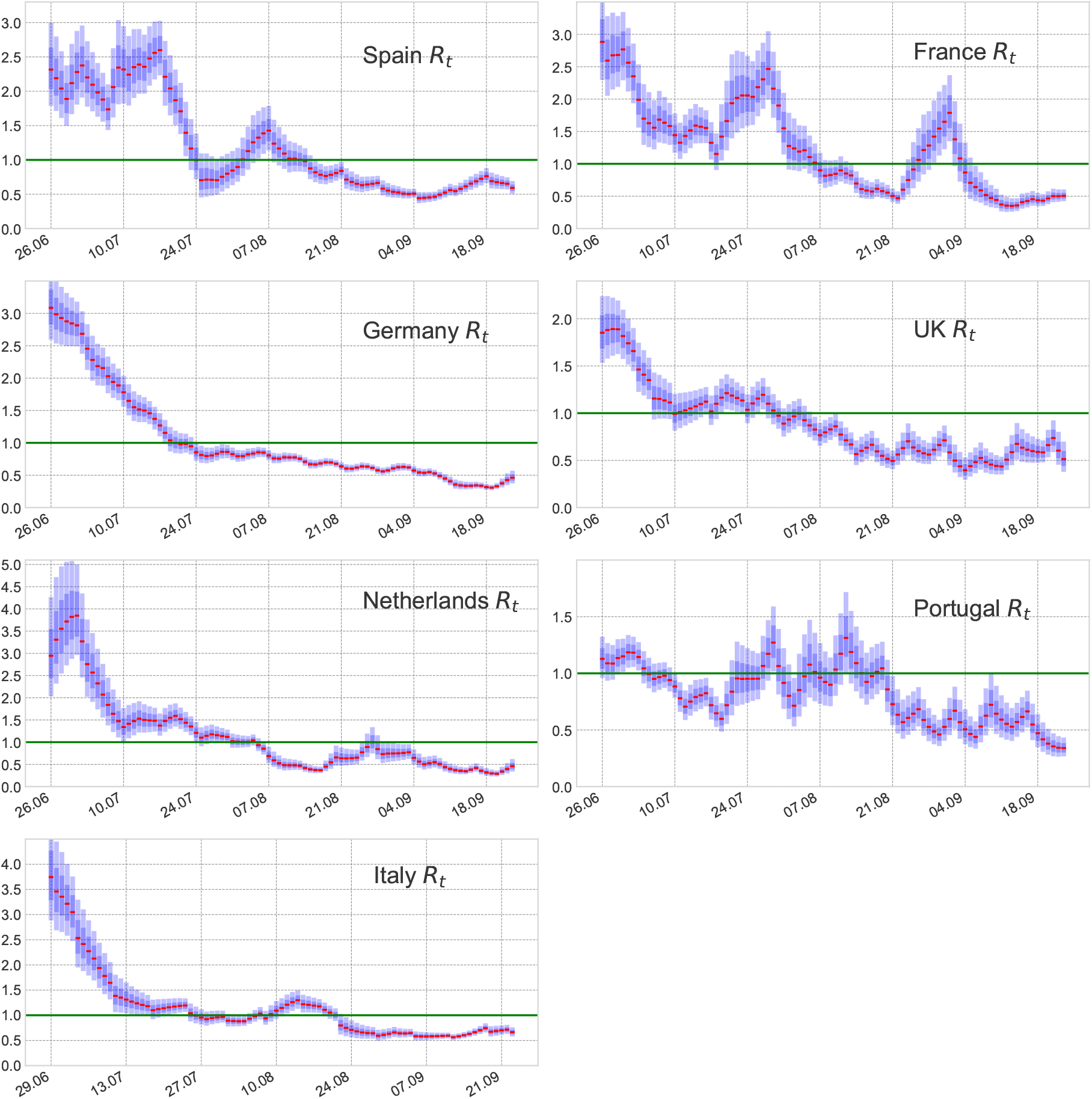
The effective reproduction number *R*_*t*_ for confirmed MPX cases in Spain, France, Germany, the UK, the Netherlands, Portugal, and Italy from June 25 to September 23, 2022. The assumed generation time distribution for the computation of *R*_*t*_ is a *Gamma*(25, 0.5) distribution. The red solid line represents the median estimate for *R*_*t*_. The dark and light blue shaded areas represent 50% and 90% quantiles for *R*_*t*_, respectively. The green solid line indicates the threshold value 1 on the reproduction number.

Observe (see Figure 4) that although Italy exhibited the highest initial instantaneous reproduction number (considering cases from the last week of June), following this period their *R*_*t*_ showed a clear decreasing trend during July reaching the threshold value 1 at the end of this month. The reproduction number for Germany showed similar patterns reaching its highest value in late June followed by a rapid decrease. The instantaneous reproduction number for Spain, France, and the UK also reached its highest value in late June. On the other hand, the *R*_*t*_ for the Netherlands reached its highest value one week late, whereas the Portuguese *R*_*t*_ showed oscillations around criticality during the whole time period. On average, the instantaneous reproduction numbers for Spain and France showed a value above one (supercritical dynamics) for a longer period of time. This is supported by the fact that both countries present the highest number of cumulative cases within the region. During the end of the period (September), all the countries considered here showed an instantaneous reproduction number below 1 which is in agreement with the significant reduction in daily confirmed MPX cases within the European area European Centre for Disease Prevention and Control (2022).

### 3.2 Growth rates and subexponential dynamics

We performed the Bayesian sequential approach for parameter estimation in the GGM model (5) and its solution (6) to characterize the epidemic growth patterns of the outbreaks in the most affected European countries. The model is tested on the smoothed incidence time series presented in Figure 1. Figures 5-6 show the parameter estimation results for the parameters *p* and *r*, respectively. The red solid line represents the median estimate, whereas the dark and light blue shaded areas represent 50% and 90% quantiles, respectively. Figure 7 presents the fits of the GGM model to the smoothed cases (see Figure 1). The estimation indicates that outbreaks in all the countries considered in this study (Spain, France, Germany, the UK, the Netherlands, Portugal, and Italy) show subexponential growth with most estimates of *p* displaying mean values below 0.5 (see Figure 5). Current monkeypox outbreaks disproportionately affected the GBMSM community showing non-homogeneous mixing and clustering in contact patterns, mechanisms that have been linked to subexponential growth dynamics for several diseases Chowell et al. (2016). The growth rate *r* for most of the countries showed an average increasing pattern except for Portugal which showed an almost constant growth rate (see Figure 6). This coincides with the *R*_*t*_ for Portugal whose value was oscillating around 1 during the whole outbreak (see Figure 4).

**Figure 5:**
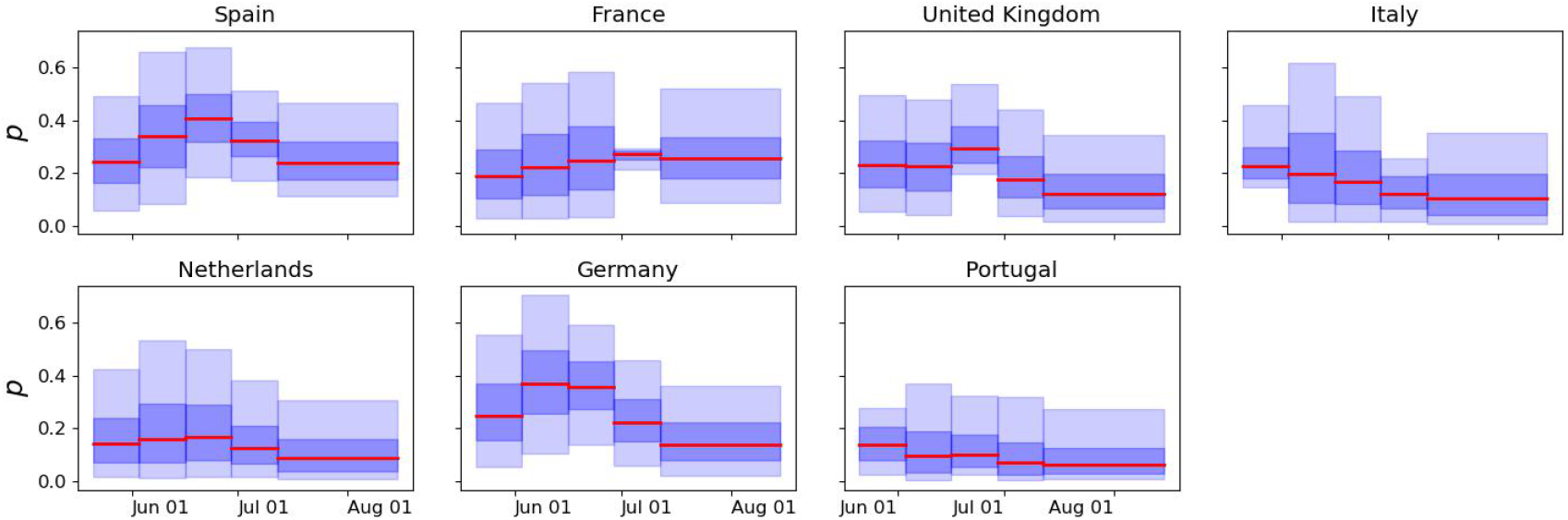
Time-varying estimates of the deceleration of growth parameter, *p*, derived from fitting the solution of the GGM model (6) to the epidemic growth phase for confirmed daily MPX cases in Spain, France, Germany, the UK, the Netherlands, Portugal, and Italy, respectively. The solid red line represents the median estimate. The dark and light blue shaded areas represent 50% and 90% quantiles, respectively.

**Figure 6:**
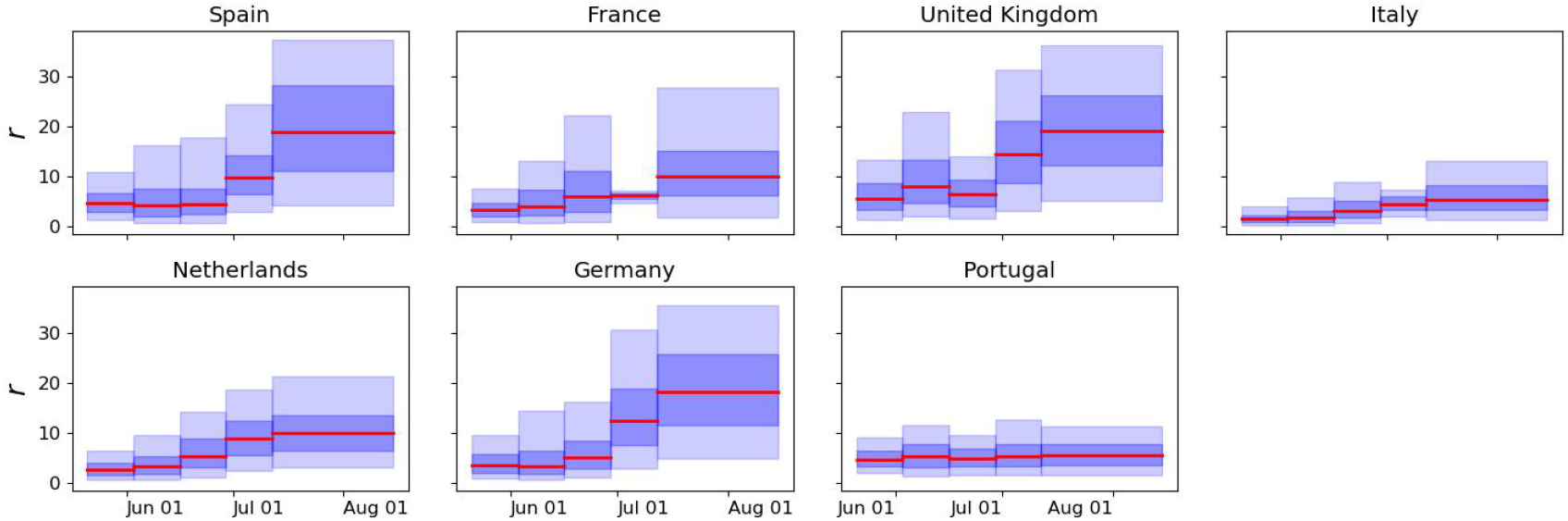
Time-varying estimates of the growth rate, *r*, derived from fitting the solution of the GGM model (6) to the epidemic growth phase for confirmed daily MPX cases in Spain, France, Germany, the UK, the Netherlands, Portugal, and Italy, respectively. The solid red line represents the median estimate. The dark and light blue shaded areas represent 50% and 90% quantiles, respectively.

**Figure 7:**
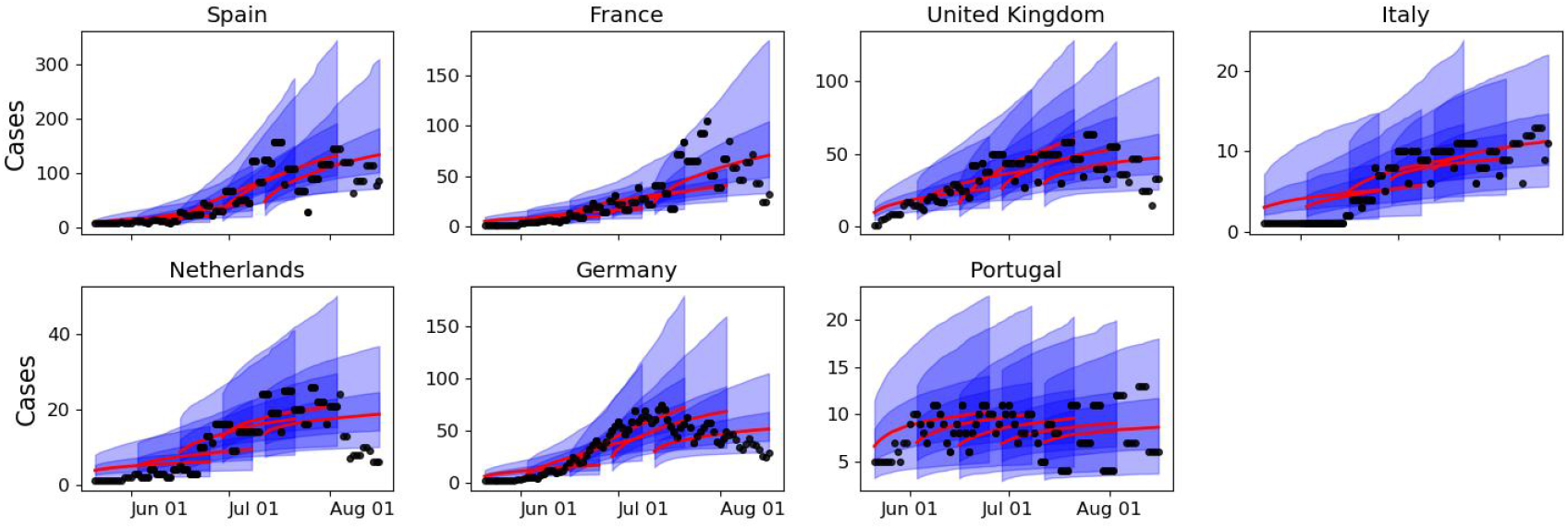
Outbreak analysis for incidence time series of MPX cases in Spain, France, Germany, the UK, the Netherlands, Portugal, and Italy. Solid red lines indicate the median incidence forecast. The darker-shaded blue region indicates the interquartile forecast range, and the lighter-shaded blue region indicates the 5–95th percentile range. Smoothed cases are presented as points.

## 4 Discussion

In this study, we have provided a reliable estimation of the instantaneous reproduction number *R*_*t*_ for the monkeypox outbreaks in the European region during June - September 2022. The estimates reported here provide useful information about pathogen transmission and can assist with outbreak control. We retrieved incidence time series on the daily laboratory-confirmed monkeypox cases available from an open-access database presented in Kraemer et al. (2022). We used a filtering type Bayesian inference that depends on a dynamic linear model on the log of observed *R*_*t*_’s to compute the instantaneous reproduction number in the European countries with the highest burden of disease in terms of cumulative cases. We have considered a plausible generation time distribution based on a recent estimation of the mean generation time in Italy Guzzetta et al. (2022). During the early phase of the outbreak, the instantaneous reproduction numbers in Spain, France, Germany, the UK, the Netherlands, Portugal, and Italy were all higher than 1 indicating supercritical epidemiological dynamics. Nevertheless, after late August most of these countries (except France) showed an *R*_*t*_ value below 1 which is in agreement with the observed reduction in monkeypox cases within the European area European Centre for Disease Prevention and Control (2022).

The current atypical monkeypox outbreaks have exposed important gaps in understanding the transmission patterns and continuously evolving epidemiological characteristics of the disease Zumla et al. (2022). Hence, we considered a simple phenomenological model that can reproduce various epidemic growth profiles with no need for explicit mechanistic assumptions about the transmission dynamics Chowell (2017). The results suggest that monkeypox outbreaks in all the countries considered in this study (Spain, France, Germany, the UK, the Netherlands, Portugal, and Italy) show subexponential growth with estimates of the deceleration of growth parameter, *p*, displaying mean values substantially lower than 1.0 (see Figure 5). Current monkeypox outbreaks disproportionately affected the GBMSM community showing non-homogeneous mixing and clustering in contact patterns, mechanisms that have been linked to subexponential growth dynamics in the past Chowell et al. (2016). Our analysis is based on a Bayesian statistical approach that allows the incorporation of background knowledge in the model’s parameters. Although our approach is well suited to model multiple sources of uncertainty, two major sources of error are worth to be mentioned. First, due to limited testing availability, especially at the early phase of the current outbreak, the daily laboratory-confirmed cases might be prone to underreporting. Second, the estimation of the instantaneous reproduction numbers is highly dependent on the generation time distribution, which has been only estimated for the case of Italy Guzzetta et al. (2022).

In summary, in this study, we report the instantaneous reproduction numbers and time-dependent estimations of the epidemic growth rates at a country level in the European region. Our findings suggest that the current monkeypox outbreaks resemble stuttering chains of transmission so the possibility of a huge outbreak is very low. Confirmed monkeypox cases are decreasing significantly in the European region. Compared to the peak of the outbreak reached during the epidemiological week 29 (18-24 July 2022), in which 2, 151 cases were reported, there has been a reduction of more than 90% in the number of newly reported cases during week 42 (17-23, October 2022) European Centre for Disease Prevention and Control (2022). The decline might be attributed to public health interventions and behavioral changes in the population due to increased risk perception. Nevertheless, further strategies toward elimination are essential to avoid the subsequent evolution of the monkeypox virus that can result in new outbreaks.

## Data Availability

The datasets analyzed during the current study are available from an open-access database presented in Kraemer, M. U., Tegally, H., Pigott, D. M., Dasgupta, A., Sheldon, J., Wilkinson, E., ... & Brownstein, J. S. (2022). Tracking the 2022 monkeypox outbreak with epidemiological data in real-time. The Lancet Infectious Diseases.

https://www.thelancet.com/journals/laninf/article/PIIS1473-3099(22)00359-0/fulltext

## Data availability

The datasets analyzed during the current study are available from an open-access database presented in Kraemer et al. (2022).

## Competing interests

The authors declare they have no known competing financial interests that could have appeared to influence the results reported in this work.

## Acknowledgments

This research is supported by the Basque Government through the “Mathematical Modeling Applied to Health” Project, BERC 2022-2025 program and by the Spanish Ministry of Sciences, Innovation and Universities: BCAM Severo Ochoa accreditation SEV-2017-0718.

## Notes

### Competing Interest Statement

The authors have declared no competing interest.

### Author Declarations

The study used only openly available human data that were originally located at: Kraemer, M. U., Tegally, H., Pigott, D. M., Dasgupta, A., Sheldon, J., Wilkinson, E., ... & Brownstein, J. S. (2022). Tracking the 2022 monkeypox outbreak with epidemiological data in real-time. The Lancet Infectious Diseases.

## References

Aguiar, M., Van-Dierdonck, J. B., and Stollenwerk, N. (2020). Reproduction ratio and growth rates: Measures for an unfolding pandemic. PLoS One, 15(7):e0236620.

Camacho, A., Saldaña, F., Barradas, I., and Jerez, S. (2019). Modeling public health campaigns for sexually transmitted infections via optimal and feedback control. Bulletin of mathematical biology, 81(10):4100–4123.

Capistrán, M. A., Capella, A., and Christen, J. A. (2022). Filtering and improved uncertainty quantification in the dynamic estimation of effective reproduction numbers. Epidemics, 40:100624.

Chowell, G. (2017). Fitting dynamic models to epidemic outbreaks with quantified uncertainty: A primer for parameter uncertainty, identifiability, and forecasts. Infectious Disease Modelling, 2(3):379–398.

Chowell, G., Sattenspiel, L., Bansal, S., and Viboud, C. (2016). Mathematical models to characterize early epidemic growth: A review. Physics of life reviews, 18:66–97.

Christen, J. A. and Fox, C. (2010). A general purpose sampling algorithm for continuous distributions (the t-walk). Bayesian Analysis, 5(2):263–282. cited By 60.

Cori, A., Ferguson, N. M., Fraser, C., and Cauchemez, S. (2013). A new framework and software to estimate time-varying reproduction numbers during epidemics. American journal of epidemiology, 178(9):1505–1512.

Daza-Torres, M. L., Capistrán, M. A., Capella, A., and Christen, J. A. (2022). Bayesian sequential data assimilation for covid-19 forecasting. Epidemics, 39:100564.

European Centre for Disease Prevention and Control (2022). Monkeypox, joint epidemiological overview. https://monkeypoxreport.ecdc.europa.eu. Accessed: 2022-11-15.

Fraser, C. (2007). Estimating individual and household reproduction numbers in an emerging epidemic. PloS one, 2(8):e758.

Guzzetta, G., Mammone, A., Ferraro, F., Caraglia, A., Rapiti, A., Marziano, V., Poletti, P., Cereda, D., Vairo, F., Mattei, G., et al. (2022). Early estimates of monkeypox incubation period, generation time, and reproduction number, italy, may–june 2022. Emerging Infectious Diseases, 28(10):2078.

Hadeler, K. (2011). Parameter identification in epidemic models. Mathematical biosciences, 229(2):185– 189.

Kraemer, M. U., Tegally, H., Pigott, D. M., Dasgupta, A., Sheldon, J., Wilkinson, E., Schultheiss, M., Han, A., Oglia, M., Marks, S., et al. (2022). Tracking the 2022 monkeypox outbreak with epidemiological data in real-time. The Lancet Infectious Diseases.

Obadia, T., Haneef, R., and Böelle, P.-Y. (2012). The r0 package: a toolbox to estimate reproduction numbers for epidemic outbreaks. BMC medical informatics and decision making, 12(1):1–9.

Parag, K., Thompson, R., and Donnelly, C. (2022). Are epidemic growth rates more informative than reproduction numbers? Journal of the Royal Statistical Society: Series A.

Poland, G. A., Kennedy, R. B., and Tosh, P. K. (2022). Prevention of monkeypox with vaccines: a rapid review. The Lancet Infectious Diseases.

Saldaña, F. and Barradas, I. (2019). The role of behavioral changes and prompt treatment in the control of stis. Infectious Disease Modelling, 4:1–10.

Saldaña, F. and Velasco-Hernández, J. X. (2021). The trade-off between mobility and vaccination for covid-19 control: a metapopulation modelling approach. Royal Society open science, 8(6):202240.

Saldaña, F. and Velasco-Hernández, J. X. (2022). Modeling the covid-19 pandemic: a primer and overview of mathematical epidemiology. SeMA Journal, 79(2):225–251.

Tsoularis, A. and Wallace, J. (2002). Analysis of logistic growth models. Mathematical biosciences, 179(1):21–55.

Viboud, C., Simonsen, L., and Chowell, G. (2016). A generalized-growth model to characterize the early ascending phase of infectious disease outbreaks. Epidemics, 15:27–37.

Wallinga, J. and Lipsitch, M. (2007). How generation intervals shape the relationship between growth rates and reproductive numbers. Proceedings of the Royal Society B: Biological Sciences, 274(1609):599–604.

Weitz, J. S., Park, S. W., Eksin, C., and Dushoff, J. (2020). Awareness-driven behavior changes can shift the shape of epidemics away from peaks and toward plateaus, shoulders, and oscillations. Proceedings of the National Academy of Sciences, 117(51):32764–32771.

World Health Organization (WHO) (2022). 2022 monkeypox outbreak: Global trends. https://worldhealthorg.shinyapps.io/mpx_global. Accessed: 2022-10-15.

Yuan, P., Tan, Y., Yang, L., Aruffo, E., Ogden, N. H., B’ selair, J., Heffernan, J., Arino, J., Watmough, J., Carabin, H., et al. (2022). Assessing transmission risks and control strategy for monkeypox as an emerging zoonosis in a metropolitan area. Journal of Medical Virology.

Zumla, A., Valdoleiros, S. R., Haider, N., Asogun, D., Ntoumi, F., Petersen, E., and Kock, R. (2022). Monkeypox outbreaks outside endemic regions: scientific and social priorities. The Lancet. Infectious Diseases.

